# Evaluating *ANO6* as a Parkinson’s disease candidate gene: a human genetic investigation of common and rare variant associations

**DOI:** 10.64898/2026.01.21.26344547

**Authors:** Sitki Cem Parlar, Hampton Leonard, Konstantin Senkevich, Lang Liu, Meron Teferra, the Global Parkinson’s Genetics Program (GP2), Ziv Gan-Or

**Author notes:** **Corresponding Author** Ziv Gan-Or, MD, PhD. **Author Emails** Sitki Cem Parlar, Hampton Leonard, Konstantin Senkevich, Lang Liu, Meron Teferra, the Global Parkinson’s Genetics Program (GP2), Ziv Gan-Or.

## Abstract

*ANO6 (TMEM16F)*, a Ca^2^□-activated lipid scramblase and ion channel, has been implicated in α-synuclein secretion and propagation, a hallmark of Parkinson’s disease (PD). To evaluate whether genetic variation in *ANO6* contributes to PD risk, we analyzed common and rare variants across large-scale datasets. Relying on genome-wide association study (GWAS) summary statistics from 63,555 PD cases, 17,700 proxy cases, and 1.7 million controls, we identified seven noncoding common variants within *ANO6* that reached genome-wide significance, but these signals were driven by linkage disequilibrium (LD) with the nearby *LRRK2* p.G2019S variant. We further tested rare variant burden in 4,879 PD cases and 65,279 controls. Rare variant burden analysis showed significant enrichment only when all rare variants were aggregated (meta-analysis *P*_*FDR*_ = 0.02), with no signal detected in functional categories. Overall, human genetic data does not support an important role for *ANO6* in PD.

---

Parkinson’s disease (PD) is a progressive movement disorder characterized by motor symptoms such as bradykinesia and tremor as well as diverse non-motor features (Poewe et al., 2017). Its etiology is complex, with genetics playing a major role in disease risk and progression (Blauwendraat et al., 2020). Over 10 genes have been implicated in PD (Blauwendraat et al., 2020), and genome-wide association studies (GWAS) have identified numerous additional loci (Kim et al., 2024; Leonard, 2025; Nalls et al., 2019). Despite these advances, much of PD heritability remains unexplained, highlighting the need to uncover additional genetic factors and pathways.

A defining pathobiological hallmark of PD is the accumulation of α-synuclein aggregates (Poewe et al., 2017). Cohen-Adiv et al. (2025) recently implicated *ANO6* (also known as TMEM16F), a Ca^2^□-activated lipid scramblase and ion channel (Suzuki et al., 2010), in this process. *ANO6* regulates phospholipid externalization and membrane dynamics and was shown to modulate extracellular α-synuclein and its neuron-to-neuron propagation (Cohen□Adiv et al., 2025). Moreover, a missense variant (p.Ala703Ser) associated with increased scramblase activity was identified in Ashkenazi Jewish PD patients (Cohen□Adiv et al., 2025).

Here, we analyze *ANO6* variation using the largest GWAS summary statistics (63,555 PD cases, 17,700 proxy cases, and 1,746,386 controls) (Leonard, 2025)to assess common variant association, and across whole-genome sequencing (WGS) datasets to test rare-variant burden, to determine whether evidence from human genetic variation converges with the recent functional evidence presented for the role of *ANO6* in α-synuclein secretion and spread (Cohen□Adiv et al., 2025).

Visualization of *ANO6* locus-wide signals was performed using LocusZoom.js (Boughton et al., 2021) (Figure 1), and summary statistics for variants within the gene region were analyzed for genome-wide significance (See Supplementary Data 1). A total of seven common variants (rs965776202, rs941040929, rs756135948, rs926284189, rs572378673, rs767368320, rs140947115) reached genome-wide significance (*P* < 5 × 10□□) for association with PD risk. Because *LRRK2* p.G2019S, a well-established PD risk variant, is located ~4.9 Mb away and previous work has shown an extended LD block of up to 7 Mb at this locus (Bar-Shira et al., 2009) we tested whether the *ANO6* association was secondary to LD with *LRRK2*. In GP2 Release 10 genotyping array data, the risk alleles at *ANO6* showed much higher frequencies in the Ashkenazi Jewish population compared with Non-Finnish Europeans from both GP2 and gnomAD (Karczewski et al., 2020) (Table 1). Using genotyping array data from GP2 Release 10, we conditioned the seven common ANO6 variants with p.G2019S in both European and Ashkenazi Jewish ancestries. After adjusting for p.G2019S, the previously observed significant associations in the *ANO6* region were no longer detected (See Supplementary Data 2), confirming that the signals were driven by LD with the *LRRK2* p.G2019S variant.

**Table 1.** Reported samples and frequencies of GWAS significant *ANO6* variants.

| Cohorts | gnomAD v4.1<br>All Populations<br>(MAF) | gnomAD v4.1<br>Non-Finnish<br>European (MAF) | GP2 R10<br>Non-Finnish<br>European (MAF) | GP2 R10<br>Ashkenazi<br>Jewish (MAF) |
| --- | --- | --- | --- | --- |
| rs965776202 | $7.21 \times 10^{-3}$ | $7.35 \times 10^{-5}$ | $1.78 \times 10^{-3}$ | 0.095 |
| rs941040929 | $2.64 \times 10^{-4}$ | $5.93 \times 10^{-5}$ | $4.45 \times 10^{-4}$ | 0.054 |
| rs756135948 | - | - | $3.05 \times 10^{-4}$ | 0.053 |
| rs926284189 | $1.65 \times 10^{-4}$ | $7.37 \times 10^{-5}$ | $3.38 \times 10^{-4}$ | 0.053 |
| rs572378673 | $7.68 \times 10^{-4}$ | $1.32 \times 10^{-4}$ | $2.90 \times 10^{-4}$ | 0.054 |
| rs767368320 | $1.91 \times 10^{-4}$ | $7.36 \times 10^{-5}$ | $2.90 \times 10^{-4}$ | 0.053 |
| rs140947115 | $8.02 \times 10^{-4}$ | $4.22 \times 10^{-4}$ | $3.40 \times 10^{-4}$ | 0.054 |

Abbreviations: MAF, minor allele frequency; gnomAD, The Genome Aggregation Database; GP2 R10, The Global Parkinson's Genetics Program Data Release 10

**Figure 1.**
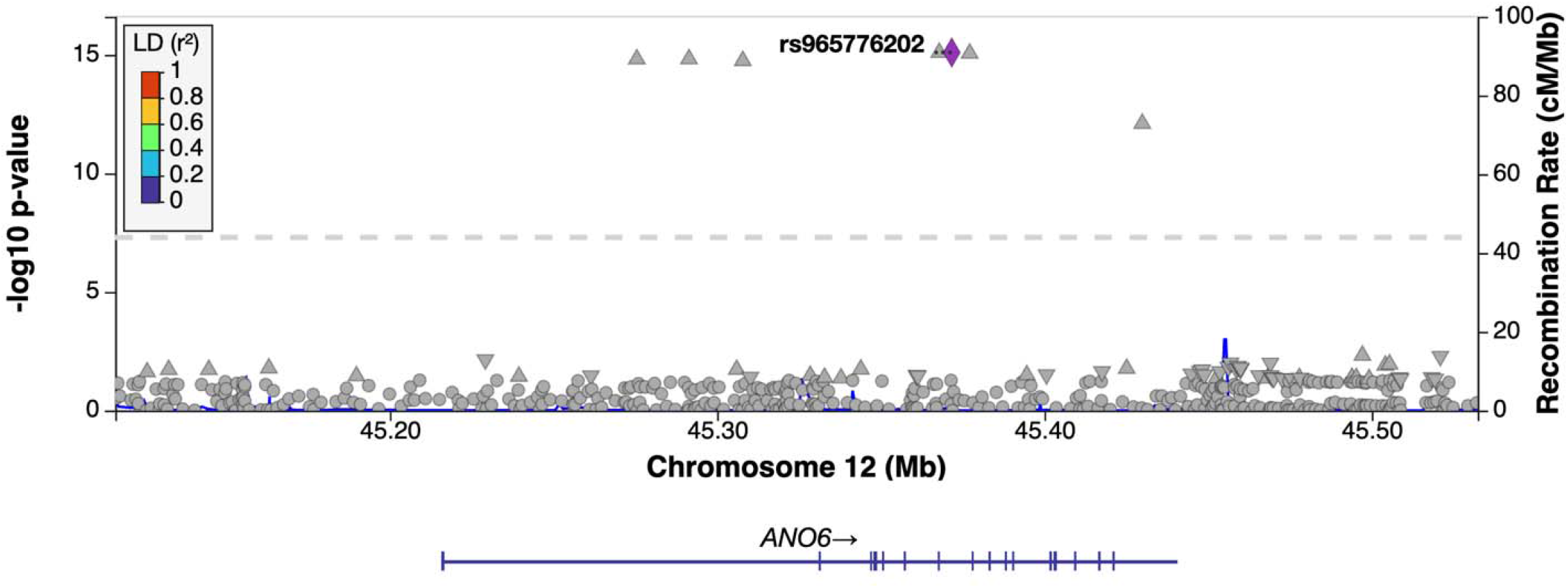
Locus zoom plot of *ANO6* (± 100kb) in Parkinson’s Disease GWAS. The x-axis represents genomic position on chromosome 12 (Mb), and the left y-axis shows the – log□□(p-value) for association with Parkinson’s Disease from the largest PD GWAS to date. Each point represents a common variant within the *ANO6* region, with color indicating linkage disequilibrium (LD, r^2^) with the lead variant (rs965776202), which is displayed as a purple diamond. The dashed horizontal line indicates the threshold for genome-wide significance (*P-value* = 5 × 10□□). The right y-axis shows the local recombination rate in centimorgans per megabase (cM/Mb), plotted as a blue line. The *ANO6* gene structure is shown at the bottom, with exons represented by vertical lines and the arrow indicating transcriptional direction.

We conducted a rare variant burden analysis using quality-controlled WGS data from two cohorts: UK Biobank (UKBB) (Consortium, 2025) and The Accelerating Medicines Partnership program for Parkinson’s disease (AMP-PD) (Iwaki et al., 2021). In total, the analysis included 4,879 PD cases and 65,279 controls (further detailed in Supplementary Data 3) and 12,144 rare variants in the *ANO6* gene. Annotated variant lists for each cohort are provided in Supplementary Data 4 and 5 for UKBB and AMP-PD respectively.

Rare variant burden was analyzed across five variant categories: all rare, non-synonymous, CADD ≥ 20, AlphaMissense predicted likely pathogenic, and loss-of-function. We calculated rare variant burden using the optimized sequence kernel association test (SKAT-O) (Lee et al., 2012) at the cohort level, followed by meta-analysis using MetaSKAT (Lee et al., 2013). After applying false discovery rate (FDR) corrections (Benjamini & Hochberg, 1995), we observed a significant association for all rare variants group in both AMP-PD (*P*_*FDR*_ = 0.02) and the meta-analysis (*P*_*FDR*_ = 0.02). However, no significance was observed in the other four categories. This suggests that the observed significant association appears to be driven by noncoding variants. Additionally, we ran the analysis adjusted for *LRRK2* p.G2019S carrier status. The adjustment did not significantly alter the outcomes, suggesting that the rare variant burden was not p.G2019S dependent. Complete SKAT-O cohort-level results and MetaSKAT meta-analysis results are available in Supplementary Data 6 and 7.

The previously reported missense variant p.Ala703Ser, which increases scramblase activity (Cohen□Adiv et al., 2025), was observed in AMP-PD (2 cases, 4 controls) but not UKBB. However, no enrichment was evident based on case-control counts, and no significance was observed in nonsynonymous burden testing either at the cohort or meta-analysis level.

Our study has several limitations. Our rare variant burden analyses were limited to individuals of European ancestry, whereas the common variant GWAS meta-analyses also included additional populations of primarily European descent, such as Finnish, Icelandic, and Ashkenazi Jewish cohorts. This limits the generalizability of our findings and may neglect population-specific variants associated with PD in other ancestries. Second, although the UKBB and AMP-PD WGS datasets were both subject to strict quality control procedures, these protocols differed between cohorts, introducing potential variability. Third, while recent GWASs in non-European populations did not identify *ANO6* as a risk locus, this may reflect the smaller sample sizes rather than true absence of association (Foo et al., 2020; Kim et al., 2024; Rizig et al., 2023). Larger and more diverse cohorts will be needed to clarify the role of *ANO6* across non-European populations.

Overall, the association between *ANO6* and PD is driven by LD with the established *LRRK2* risk variant p.G2019S. The rare variant burden appears limited to noncoding association and is potentially spurious. Thus, our study suggests that *ANO6* has minor or no role in PD.

## METHODS

### Common variant analysis

LocusZoom.js was used to visualize the common variant associations and pinpoint potentially causal variants (Boughton et al., 2021). To assess whether the *ANO6* association signal was independent of the known *LRRK2* p.G2019S variant, we conditioned common ANO6 variants with p.G2019S using the –condition flag in Plink2 (Purcell et al., 2007). Conditioning was performed using 1,813 cases and 996 controls of Ashkenazi Jewish ancestry and 26,358 cases and 20,088 controls of Non-Finnish European ancestry from GP2 Release 10.

All GP2 samples used for the conditioning analyses (release 10; DOI 10.5281/zenodo.15748014) were genotyped on the Illumina NeuroBooster Array (Bandres-Ciga et al. 2023). Raw array data were processed using the GenoTools pipeline (available at https://github.com/GP2code/GenoTools; Koretsky et al. 2022), which incorporates a machine-learning framework for ancestry prediction and relatedness pruning. Sample-level QC excluded individuals with call rate <95%, discordance between genetic and reported sex, excess heterozygosity (|F|>0.25), or relatedness exceeding 12.5% IBD (approximately 2nd degree relatives). SNP-level QC removed variants with SNP-level missingness >5%, HWE P<1E-4 in controls, or with differential missingness by case-control status or haplotype at P<=1E-4. QC’d data was imputed per ancestry group using the TOPMed r3 reference panel.

### Rare variant analysis

We analyzed 4,779 PD cases and 65,525 controls from two large WGS cohorts: UKBB and AMP-PD. Whole-genome sequencing was performed using the Illumina NovaSeq 6000 S4 (UKBB) (Consortium, 2025) and HiSeq X Ten (AMP-PD) (Iwaki et al., 2021) platforms, aligned to hg38. ANO6 variants (chr12:45,216,322–45,429,308; UniProt Q4KMQ2-1) (“UniProt: the universal protein knowledgebase in 2025,” 2025) were extracted, retaining only unrelated European ancestry samples. PD diagnosis followed UK Brain or Movement Disorders criteria (Hughes et al., 1992; Postuma et al., 2015).

Quality control steps were applied in both cohorts. In UKBB, multi-allelic variants were removed using bcftools (Danecek et al., 2021), and quality control filters were applied using Genome Analysis Toolkit (GATK) v4.2.5 (Van der Auwera & O’Connor, 2020), retaining only high-quality data by setting genotype quality (GQ) ≥ 25, read depth (DP) ≥ 25, and excluding variants with >5% missingness. In AMP-PD, the data came readily quality controlled as per the steps described in their website (https://amp-pd.org/whole-genome-data) as well as their published article (Iwaki et al., 2021). Finally, only rare variants were retained via minor allele frequency (MAF) < 0.01 filter.

For functional categorization, variant annotation was performed using the Ensembl Variant Annotation Predictor (VEP) annotation software v107.0 (McLaren et al., 2016) based on GENCODE V45 (Mudge et al., 2025). The rare variants were scored based on predicted deleteriousness via Combined Annotation Dependent Depletion (CADD) v1.7 (Schubach et al., 2024) and predicted pathogenicity via AlphaMissense (Cheng et al., 2023). Further, per-variant allele frequencies across different populations were extracted from The Genome Aggregation Database (gnomAD) (Karczewski et al., 2020).

*ANO6* rare variants were analyzed for their burden for PD risk in five groups: (1) all rare variants, (2) all non-synonymous coding variants, (3) CADD≥20 (top 1% of predicted deleterious) variants, (4) AlphaMissense predicted likely pathogenic variants, (5) loss-of-function variants (stopgain, frameshift and splice-site (±1 and ±2). The analysis was conducted using the SKAT-O (Lee et al., 2012) and MetaSKAT (Lee et al., 2013) R-packages for the cohort-level analysis and meta-analysis respectively. We adjusted for sex and age as covariates to control for their potential confounding effects. Lastly, multiple-test corrections were applied using the FDR method to minimize false-positive findings (Benjamini & Hochberg, 1995). A secondary analysis was done also adjusting for *LRRK2* p.G2019S carrier status.

## ACKNOWLEDGMENTS

We would like to express our gratitude to the participants from various cohorts for their contributions to this study. This research was conducted using the NeuroHub infrastructure and was supported in part by funding from the Canada First Research Excellence Fund, awarded through the Healthy Brains, Healthy Lives initiative at McGill University, as well as by Calcul Québec and the Digital Research Alliance of Canada. Additionally, the study utilized data from the UK Biobank under Application Number 45551. Data for this article were also obtained in January 2023 from the Accelerating Medicines Partnership® (AMP®) Parkinson’s Disease (AMP PD) Knowledge Platform as per release 2.5. Since this release more recent versions have been made available. For the most up-to-date information on AMP-PD, please visit https://www.amp-pd.org. The AMP® PD program operates as a public-private partnership managed by the Foundation for the National Institutes of Health (FNIH) and funded by the National Institute of Neurological Disorders and Stroke (NINDS) in collaboration with the Aligning Science Across Parkinson’s (ASAP) initiative. Industry partners include Celgene Corporation (a subsidiary of Bristol-Myers Squibb Company), GlaxoSmithKline plc (GSK), The Michael J. Fox Foundation for Parkinson’s Research (MJFF), Pfizer Inc., AbbVie Inc., Sanofi US Services Inc., and Verily Life Sciences. “ACCELERATING MEDICINES PARTNERSHIP” and “AMP” are registered service marks of the U.S. Department of Health and Human Services. The clinical data and biosamples utilized for this article were derived from the following cohorts: (i) the BioFIND study (MJFF and NINDS); (ii) the Harvard Biomarkers Study (HBS) and the Stephen & Denise Adams Center for Parkinson’s Disease Research at Yale School of Medicine (CPDR-Y); (iii) the National Institute on Aging (NIA) International Lewy Body Dementia Genetics Consortium Genome Sequencing in Lewy Body Dementia Case-control Cohort (LBD); (iv) the MJFF LRRK2 Cohort Consortium (LCC); (v) the NINDS Parkinson’s Disease Biomarkers Program (PDBP); (vi) the MJFF Parkinson’s Progression Markers Initiative (PPMI); (vii) the NINDS Study of Isradipine as a Disease-modifying Agent in Subjects With Early Parkinson Disease, Phase 3 (STEADY-PD3); and (viii) the NINDS Study of Urate Elevation in Parkinson’s Disease, Phase 3 (SURE-PD3). BioFIND is sponsored by MJFF with support from NINDS; up-to-date study information is available at michaeljfox.org/news/biofind. The BioFIND investigators did not participate in the review of this manuscript’s data analysis or content. Genome sequence data for the Lewy body dementia case-control cohort were generated at the U.S. National Institutes of Health Intramural Research Program, supported in part by the NIA (program #: 1ZIAAG000935) and NINDS (program #: 1ZIANS003154). The Harvard Biomarker Study (HBS) is a collaboration of HBS investigators (listed at https://www.bwhparkinsoncenter.org/biobank/) and is funded through philanthropy, NIH, and non-NIH sources. The Stephen & Denise Adams Center for Parkinson’s Disease Research of Yale School of Medicine is similarly funded through philanthropy, NIH, and non-NIH sources. Neither the HBS nor CPDR-Y investigators participated in reviewing the data analysis or content of this manuscript. Data were obtained from the MJFF-sponsored LRRK2 Cohort Consortium (LCC). Current study information can be found at https://www.michaeljfox.org/biospecimens. The LCC investigators did not review the data analysis or manuscript content. PPMI is sponsored by MJFF and supported by a consortium of scientific partners (full list available at https://www.ppmi-info.org/about-ppmi/who-we-are/study-sponsors). For up-to-date study information, visit www.ppmi-info.org. PPMI investigators did not participate in the review of the data analysis or content of this manuscript. The Parkinson’s Disease Biomarker Program (PDBP) consortium is supported by NINDS at the NIH. A full list of PDBP investigators is available at https://pdbp.ninds.nih.gov/policy. PDBP investigators did not review the data analysis or content of this manuscript. STEADY-PD3 is funded by NINDS at the NIH with support from MJFF and the Parkinson Study Group. Additional information is available at https://clinicaltrials.gov/ct2/show/study/NCT02168842. The STEADY-PD3 investigators did not review the data analysis or content of this manuscript. SURE-PD3 is funded by NINDS at the NIH with support from MJFF and the Parkinson Study Group. Additional information is available at https://clinicaltrials.gov/ct2/show/NCT02642393. The SURE-PD3 investigators did not review the data analysis or content of this manuscript.

Data (DOI 10.5281/zenodo.15748014, release 10) and/or code used in the preparation of this article were obtained from The Global Parkinson’s Genetics Program (GP2). GP2 is funded by the Aligning Science Across Parkinson’s (ASAP) initiative and implemented by The Michael J. Fox Foundation for Parkinson’s Research (https://gp2.org). For a complete list of GP2 members see https://gp2.org.

## AUTHOR CONTRIBUTIONS

S.C.P. led the conception, planning, and implementation of the research project, including the design, data acquisition, analysis, and preparation of the initial manuscript draft. H.L. contributed to data acquisition and analysis. K.S. was involved in analysis and provided critical revisions.L.L. contributed to the data acquisition. Z.G.-O. was involved in shaping the research concept and design of the analyses and provided critical revisions. All authors reviewed and approved the final version of the manuscript.

## DATA AVAILABILITY STATEMENT

The summary GWAS statistics used for the common variant analysis is accessible publicly. The datasets used in this study include those from the AMP-PD Knowledge Platform (https://www.amp-pd.org) and the UKBB, both of which require institutional approvals for access. All data directly relevant to this study’s analysis, including the list of analyzed variants and demographics of the study population, are provided in the supplementary files of this article. The scripts used can be accessed through the online GitHub repository at the following link: https://github.com/gan-orlab/ANO6.

